# Modelling axonal loss caused by chronic demyelination and slow-burning inflammation at the rim of chronic lesions in MS patients

**DOI:** 10.1101/2023.05.29.23290663

**Authors:** Samuel Klistorner, Alexander Klistorner

## Abstract

**Objectives:** Slow-burning inflammation at the edge of chronic multiple sclerosis lesions and loss of myelin in the depths of the lesions have emerged as a key components of disease progression. However, their relative contribution to progressive axonal damage has not been investigated. Therefore, the aim of the study was to examine relative weight of those factors in axonal attrition inside the chronic MS lesions by measuring tissue rarefication of the lesion core.

**Methods:** Pre- and post-gadolinium 3D-T1, 3D FLAIR, diffusion tensor images, Optical Coherence tomography and multifocal visual evoked potentials were acquired from 52 patients. Analysis was performed between baseline and 48 months. Lesion expansion was measured using in-house software. The degree of lesional tissue damage was determined by measuring increase of Mean Diffusivity (MD) in lesion core normalised over MD dynamic range.

**Results:** There were 104 expanding and 257 stable lesions. Rate of normalised MD (nMD) increase was several folds higher in expanding vs stable lesions (0.21% vs 1.12% per year, p=0.01). The magnitude of nMD change was significantly associated with the rate of lesion expansion (r=0.4, p<0.001). Analysis of visual system revealed the rate of axonal loss similar to the degree of tissue rarefication in stable lesions.

**Interpretation:** The current study demonstrated a significant increase in water content in chronic MS lesions, which was, however, markedly higher in slowly expanding compared to stable lesions. This suggests that slow-burning inflammation at the lesion rim, when present, is likely to play a more significant role in axonal attrition than chronic demyelination.

## Introduction

Multiple sclerosis (MS) is a complex disease of the central nervous system (CNS), characterized by inflammation, demyelination, neuroaxonal loss and gliosis. Inflammatory demyelinating lesions are a hallmark of the disease. However, neuroaxonal loss is believed to underpin the progressive disability that characterizes MS. While acute lesional damage is a major cause of axonal loss in MS, slow-burning inflammation at the edge of chronic lesions and loss of trophic support from myelin in the depths of the demyelinated lesions have also been suggested to contribute to progressive axonal damage. [1][2]

These comparatively slow processes might be concealed in the early stages of multiple sclerosis (MS) due to the presence of acute clinical symptoms accompanying the formation of new focal lesions, as well as the brain’s remarkable compensatory capacity, but are likely to become progressively more evident during the later stages of the disease. Although it is reasonable to hypothesize that both mechanisms operate concurrently and independently, their respective contributions to axonal damage remain uncertain.

Axonal transection caused by slow-burning inflammation at the rim of chronic lesions results in Wallerian and retrograde degeneration and subsequent elimination of transected axons. This, in turn, leads to rarefication of white matter tissue in corresponding lesions, as measured by an increase of isotropic water diffusion (Mean Diffusivity, MD).[3] Accelerated axon attrition caused by permanent demyelination is also likely to produce a similar effect on tissue density inside the chronic lesions. [1][4]

Nevertheless, considering the total absence of myelin in MS lesions, the harmful impact of persistent demyelination is likely to affect all chronic lesions uniformly. [1][4] However, the degree of axonal injury caused by low-grade inflammation depends on the level of chronic inflammatory activity in each lesion. Consequently, the extent of damage may vary between lesions.

Therefore, in the current study we investigated the relative contribution of permanent demyelination and slow-burning inflammation at the lesion rim on axonal attrition inside the chronic MS lesions by measuring tissue rarefication of the lesion core. Furthermore, using the visual system as a model, the effect of chronic demyelination on acceleration of axonal loss was further verified in a sub-group of patients with unilateral optic neuritis.

## Method

The study was approved by University of Sydney and Macquarie University Human Research Ethics Committees and followed the tenets of the Declaration of Helsinki. Written informed consent was obtained from all participants.

### Subjects

Consecutive RRMS patients diagnosed according to the revised McDonald 2017 criteria[5] who were enrolled in longitudinal study of MS-related axonal loss and who completed at least 5 years follow-up were included in the study.

Patients underwent MRI scans, Optical Coherence Tomography (OCT), Multi-focal Visual Evoked potentials (mfVEP) and clinical assessment annually.

### MRI analysis

The main MRI analysis was performed between 12 months (termed “baseline”) and 60 months (termed “4 years follow-up”), while scans performed at 0 months (termed “pre-study scans”) were used to identify (and exclude) newly developed lesions at the start of the study (i.e. between 0 and 12 months).[6]

### MRI protocol and analysis

MRI was performed using a 3T GE Discovery MR750 scanner (GE Medical Systems, Milwaukee, WI). The following MRI sequences were acquired: Pre- and post-contrast (gadolinium) sagittal 3D T1, FLAIR CUBE and diffusion weighted MRI. The specific acquisition parameters and MRI image processing are provided in the **Supplementary material**.

JIM 9 software (Xinapse Systems, Essex, UK) was used to segment individual lesions at baseline and 4 years of follow-up (using co-registered T2 FLAIR images). Lesional activity during those 48 months was analysed using custom-built software that identifies the stable and expanding components of chronic lesions (adjusted to correct for brain atrophy-related displacement of lesions at follow-up [6]) and new lesions, as described previously.[7]

Central brain atrophy (CBA) was measured by calculating the volumetric change of the lateral ventricles between baseline and follow-up.[8][9] [10] This was determined by multiplying baseline ventricle volume (as segmented by SIENAX) by the percentage ventricular volume change (as calculated by VIENA, an FSL tool).[11]

In this study, the severity of slow-burning inflammation at the rim of chronic lesions was estimated by its imaging equivalent, i.e. the extent of lesion expansion. Based on the degree of expansion, chronic MS lesions were categorised into expanding and stable groups. A minimum local expansion of 4% per year was used as a cut-off for determining expanding lesions, as previously suggested[12] Lesions smaller than 100 voxel were excluded due to partial volume effect, and lesions that shrank by more than 16% were excluded from the analysis.

Lesion core was identified as the lesion volume eroded by 1 voxel from all sides.

The degree of lesional tissue damage was determined by measuring the increase in Mean Diffusivity (MD)[13] inside the lesion core between baseline and follow-up scans, as described previously [3] [14]. To quantify the measurement, the MD change was normalised (stratified) (nMD) by the dynamic range of isotropic water diffusion in the white matter, calculated based on the following assumptions:

a. MD in NAWM characterises largely preserved axonal density
b. The highest MD value detected in the lesion core across all the subjects represents a total or near-total axonal loss
c. MD gradient within the dynamic range is proportional to the level of tissue rarefication and, consequently, to the axonal loss (as demonstrated by MRI-histopathological correlation). [15]

We hypothesised that the dynamic range of MD in the white matter of MS patients incorporates an entire spectrum of the tissue damage from normal density of the brain tissue (i.e. NAWM) to extreme tissue rarefication (i.e. severe axonal loss in chronic black holes)[16] [17] and, therefore, can be calculated as a difference between the corresponding MD values, i.e. between the peak distribution of MD value in NAWM and the maximum MD value in a core of chronic lesion.

MD of NAWM was calculated as follows: the white matter mask was extracted using the brain extraction tool from ANTs (Advanced Neuroimaging Tools). The white matter mask was eroded by 1 voxel to remove partial voluming from grey matter and CSF. The mask was further reduced by removing voxels of a dilated (by 1 voxel) lesion mask. The final NAWM mask was overlaid with DTI image to calculate the MD of NAWM.

To calculate the relative (%-wise) increase of tissue rarefication, the difference in MD between follow-up and baseline timepoints in the lesion core was normalised by the MD dynamic range.

To minimize the effect of Wallerian and retrograde degeneration caused by transection of axons in newly appearing lesions on alteration of MD in chronic lesion core, the voxels containing fiber tracts passing through new lesions were removed from the analysis. This was implemented as follows: firstly, full brain tractography was generated using the TractSeg algorithm; secondly, generated tracts were intersected with new lesions at follow-up to create ‘new lesion tracts’; finally, ‘new lesion tracts’ were intersected with the core of chronic lesions and intersecting voxels were removed from the analysis (Fig.1).

**Fig. 1.**
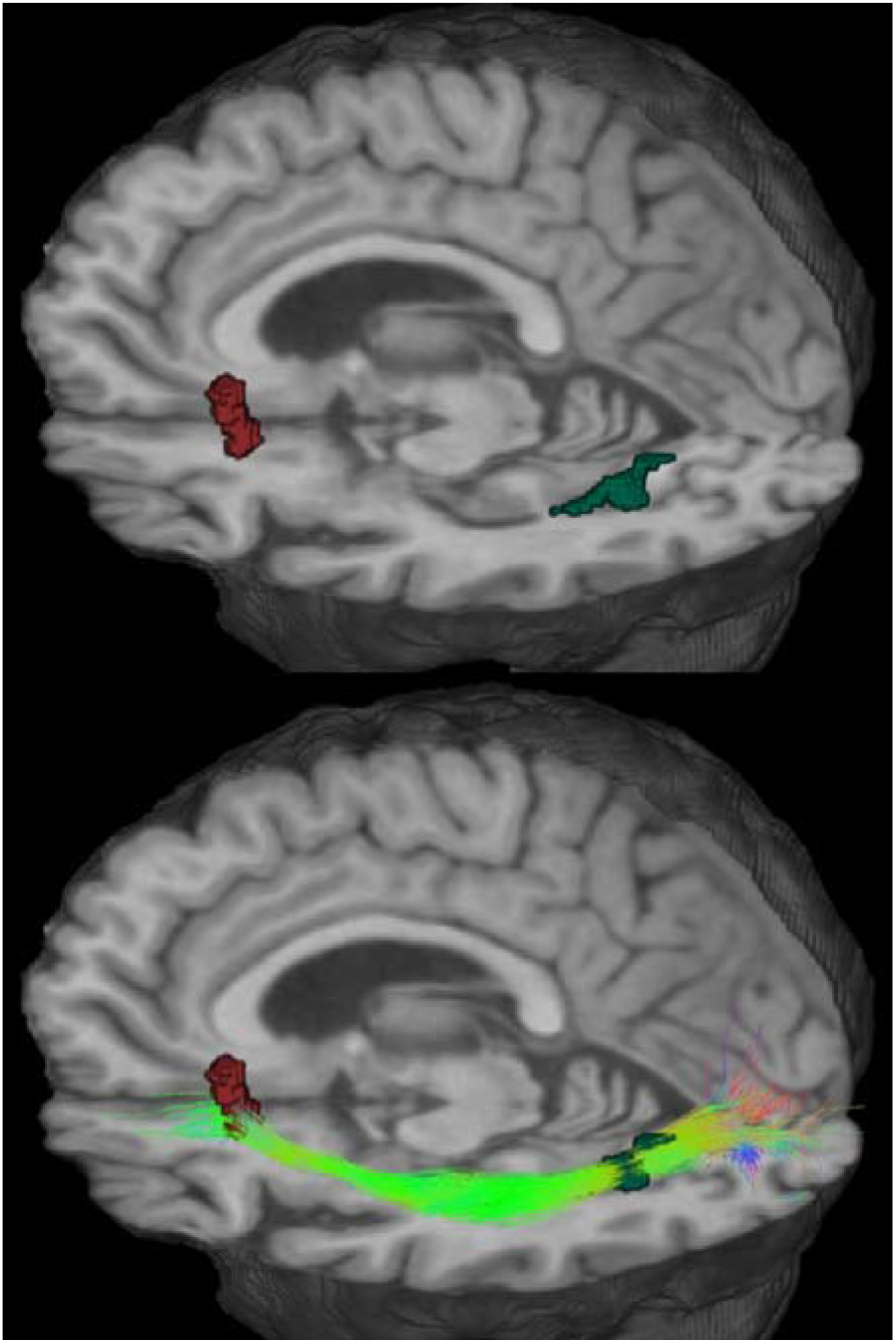
Elimination of voxels contaminated by axons potentially transected within new lesions. Top: example of new (green) and chronic (red) lesions. Bottom: tractography streamlines intersecting both new and chronic lesions.

### Analysis of the visual system

For the analysis of the visual system, the sub-group of patients with a history of unilateral ON at least 12 months prior to enrolment and a significant degree of optic nerve demyelination was identified within the main cohort. The diagnosis of ON was based on clinical findings, which included an appropriate history and objective examination findings (decreased visual acuity, a visual field defect, colour vision loss, relative afferent pupil defect, and a compatible fundus examination).

All participants underwent a peripapillary ring scan at baseline and follow-up using the Heidelberg Spectralis OCT and Eye Explorer software version 6.9.5.0 (Heidelberg Engineering, Germany) as described previously [18]. In this study, the OCT data were reported according to the APOSTEL recommendations [19]. The pupils of participants were not dilated, and the scans were performed by the same operator and using the same device under room light conditions. The peripapillary ring scan (manual placement of the ring; diameter = 3.50 mm) was used to obtain retinal nerve fiber layer (RNFL) thickness measures. Global RNFL thickness was analysed. For subsequent visits, the follow-up function was activated when scans were performed to ensure that they were obtained at the same locations as the baseline scans. All images were checked, and those with poor quality and segmentation errors were excluded to ensure that all OCT images included in the analysis fulfilled the OSCAR-IB criteria [20]. Similar to MD, analysis of RNFL thinning in both ON and fellow eyes was compared to the baseline RNFL value in the fellow eye.

The level of optic nerve demyelination was assessed by the latency delay of mfVEP recorded using the Vision Search system (VisionSearch, Sydney, Australia) with standard stimulus conditions as described previously [21]. In brief, four gold-disc electrodes (Grass, West Warwick, RI) were used for bipolar recording with two electrodes positioned 4 cm on either side of the inion, one electrode 2.5 cm above and another 4.5 cm below the inion in the midline. Electrical signals were recorded along two channels, measured as the difference between superior and inferior and between the left and right electrodes. The quality of the VEP signal was assessed by AI-assisted in-house software, which was also used to estimate the latency of individual segments. [22] Since post-chiasmal/optic radiation lesions are likely to have similar effects on latency delay in both eyes and in order to reduce between-subject variability, inter-eye asymmetry of the mfVEP latency was used to determine the level of ON-related demyelination in the affected eye.

### Statistical analysis

Statistical analysis was performed using SPSS 22.0 (SPSS, Chicago, IL, USA). Pearson correlation coefficient was used to measure statistical dependence between two numerical variables. P < 0.05 was considered statistically significant. Shapiro–Wilk test was used to test for normal distribution. Longitudinal changes were assessed using a paired two-sample t-test. To investigate how well each factor predicts central brain atrophy, we performed a forward stepwise linear regression analysis that included age, gender and disease duration.

## Results

Fifty-two consecutive MS patients who were enrolled in a longitudinal study of MS-related axonal loss and who had 5 years of follow-up satisfied the inclusion criteria and were included in the study. Nine patients were maintained on low-efficacy treatment (injectables, such as interferon and glatiramer acetate, teriflunomide and dimethyl-fumarate)[23], while 25 patients were receiving high-efficacy drugs (fingolimod, natalizumab, and alemtuzumab)[23] during the study period. Three patients were treatment-free, while 15 patients changed treatment category between baseline and follow-up visits. Demographic data is presented in Table 1.

**Table 1.** Demographic data for study patients.

|  | RRMS patients |
| --- | --- |
| N | 52 |
| Age, years (Mean±SD) | 41.7 ± 9.1 |
| F/M ratio | 1.6/1 |
| Disease duration, years (Mean±SD) | 5.5 ± 4.1 |
| EDSS at baseline (median (range)) | 1 (0-4) |

Fifteen patients from the study cohort had a history of unilateral ON and demonstrated a significant degree of demyelination in ON eyes with a minimum inter-eye mfVEP latency delay of 15 ms (indicating that at least one-third of the optic nerve length was demyelinated). The average inter-eye latency asymmetry of mfVEP in this group was 26.3+/-9.8 ms

### MD dynamic range

The average MD value in voxels of NAWM combined from all subjects was 0.76+/-0.09 μm2/ms. The distribution of NAWM MD values was very tight (coefficient of variability-11.8%) (Fig 3, blue graph). Conversely, the distribution of MD in voxels restricted to lesion core was noticeably broader, reflecting the wide spectrum of lesional tissue damage[24]. The highest (99 percentile) value of MD in lesion core across all the subjects (examples presented in Fig.2), which is likely to indicate very extensive, or potentially total, axonal loss, reached 1.95 μm^2^/ms) (Fig 3, orange graph). Therefore, the calculated dynamic range of MD (i.e. range between fully destroyed tissue and normal density tissue) was 1.19 μm^2^/ms (1.95 μm^2^/ms - 0.76 μm^2^/ms) (Fig 3). This value was used to normalise MD (nMD) change during follow-up in order to quantify tissue damage in the lesion core.

**Fig. 2.**
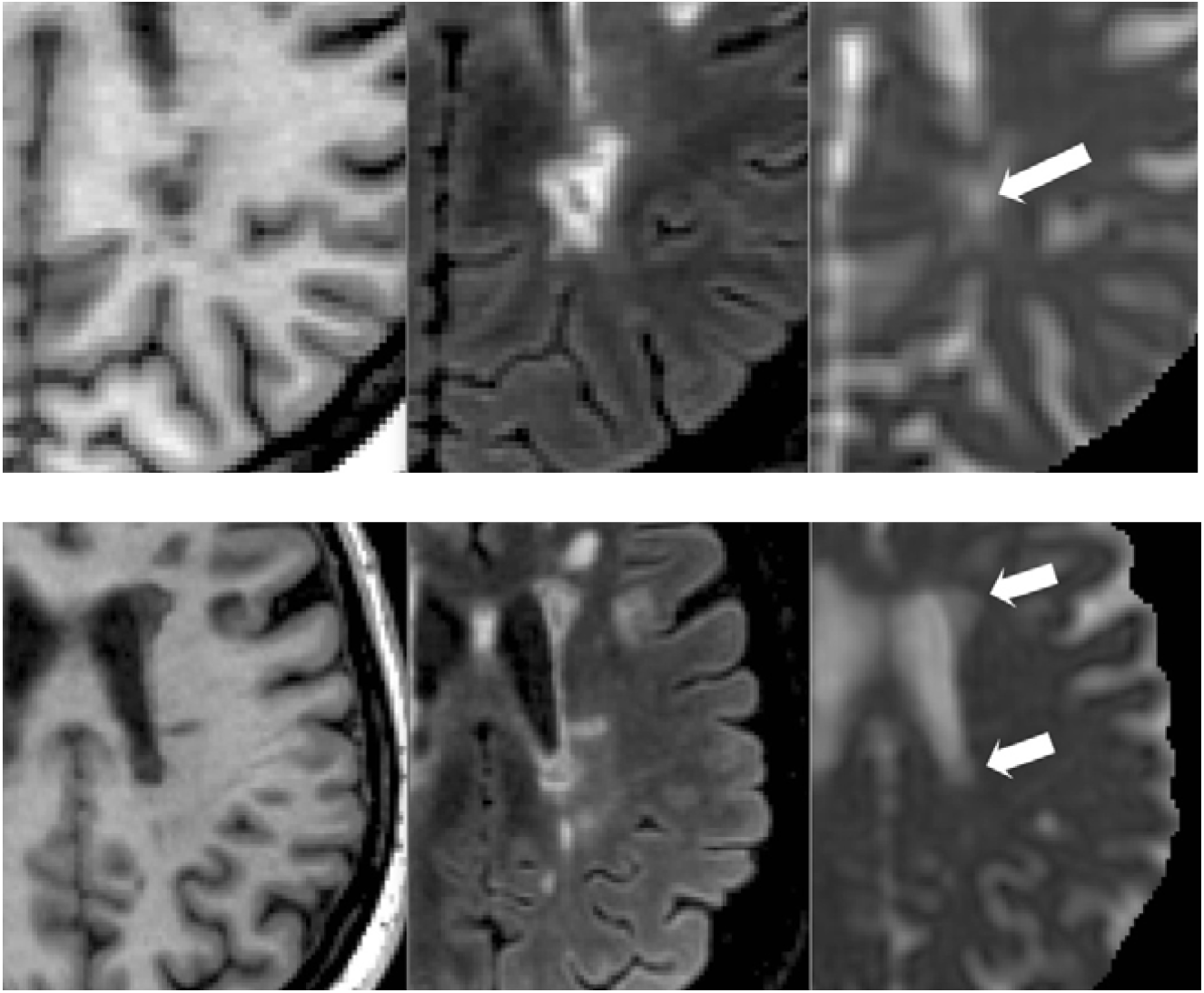
Destructive chronic MS lesion, so called “black hole” (arrows). T1 (left), FLAIR (center) and MD (right) images. Highest value of MD in the lesion core reached 1.93 μm^2^/ms in one patient (top row) and 1.95 μm^2^/ms-in another patient (bottom row).

**Fig. 3.**
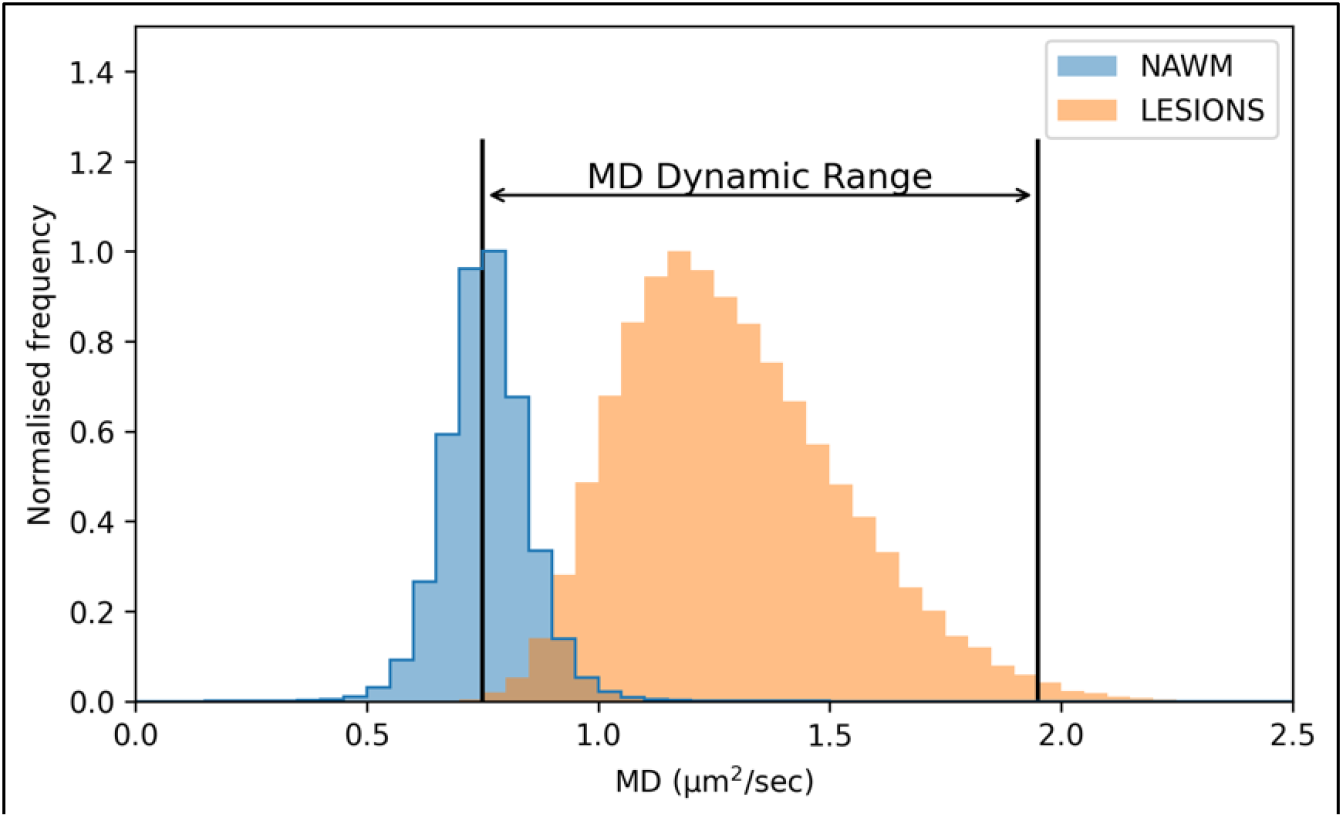
Histogram of MD distribution in voxels of NAWM (blue) and chronic lesions (orange). MD dynamic range was calculated as difference between average MD value in NAWM and 99^th^ percentile MD value in lesions.

### Progressive rarefication of tissue in chronic brain MS lesions

361 lesions were analysed in this study. Based on mean lesional MD at baseline (Table 2) and follow the rationale described in previous section, the average degree of tissue rarefication in the core of chronic lesions as compared to NAWM (i.e. extent of tissue loss) was 37%. {(1.2-0.76)/1.19}.

**Table 2.**

| | Baseline volume | Follow-up volume | p-value Paired ttest) | Baseline MD $\mu\text{m}^2/\text{ms}$ | Follow-up MD $\mu\text{m}^2/\text{ms}$ | p-value Paired ttest) |
| --- | --- | --- | --- | --- | --- | --- |
| Stable | 723±1376 | 783±1536 | <0.001 | 1.20±0.18 | 1.21±0.19 | 0.014 |
| Expanding | 615±789 | 845±1043 | <0.001 | 1.24±0.17 | 1.29±0.19 | <0.001 |

There were 104 expanding and 257 stable lesions. Proportion of stable and expanding lesions in individual patients is presented in Figure 4. Thirty-six patients had at least one expanding lesion.

**Fig. 4.**
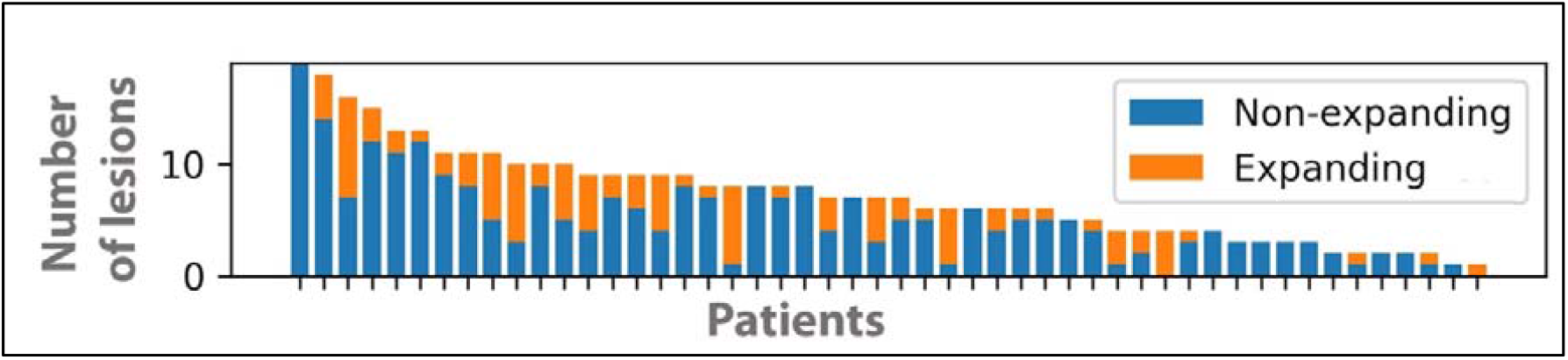
Subject-wise distribution of stable and expanding lesions.

Baseline and follow-up average volume of chronic lesions and MD values are presented in Table 2

The average increase of stable lesion volume was 8%, while expanding lesions increased in size on average by 37.4%. Both expanding and stable lesions demonstrated a significant degree of MD increase in the lesion core during follow-up (Table 2), which was, however, markedly larger in expanding lesions (p <0.0001 and p=0.04 for expanding and stable lesions respectively, paired t-test).

Accordingly, analysis of stable lesions revealed a 0.85% increase of nMD during the follow-up time (4 years) or 0.21% per year. In contrast, in chronic expanding MS lesions, the proportion of isotropic water diffusion increased on average by 4.8% during 4 years of follow-up or 1.2% per year. Furthermore, the magnitude of nMD change was significantly associated with the rate of lesion expansion (r=0.4, p<0.001) (Fig. 5). This correlation remained significant when limited to expanding group only (r=0.3, p=0.002). Note that relative (percentagewise) MD change is normalised based on MD dynamic range.

**Fig. 5.**
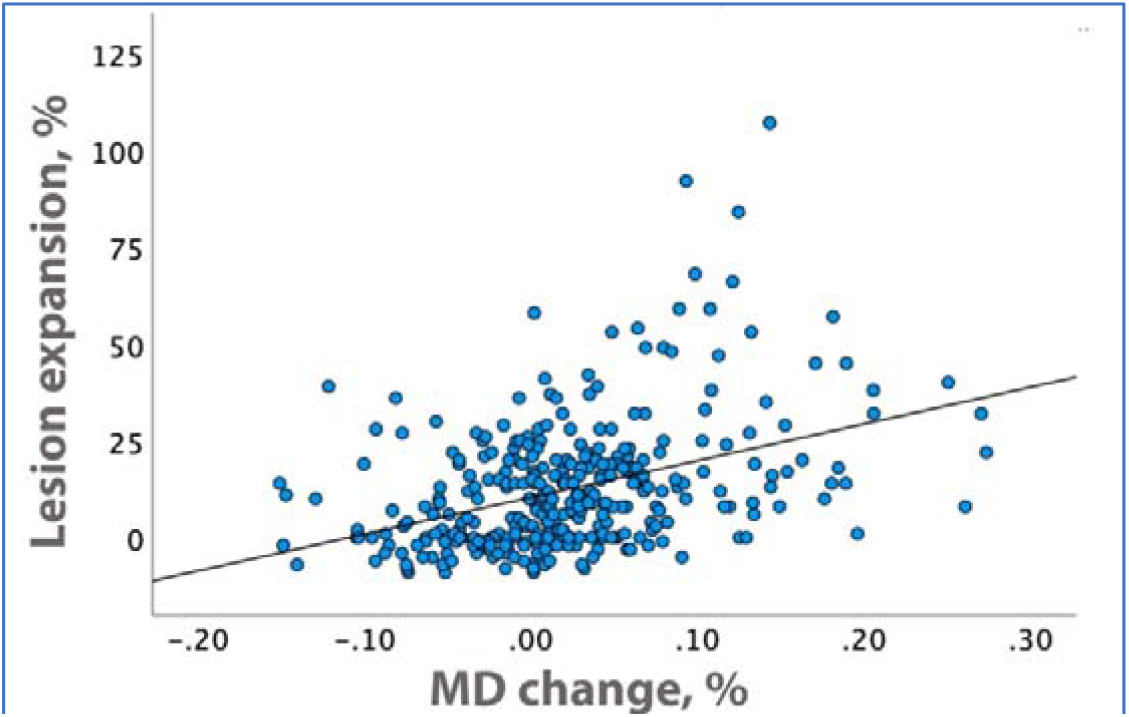
Correlation between nMD change in lesion core and the rate of chronic lesion expansion.

Figure 6a, which represents a normalised distribution of nMD change in individual voxels of the lesion core in stable and expanding lesion groups, shows a noticeable shift of expanding lesions toward higher MD values.

**Fig. 6.**
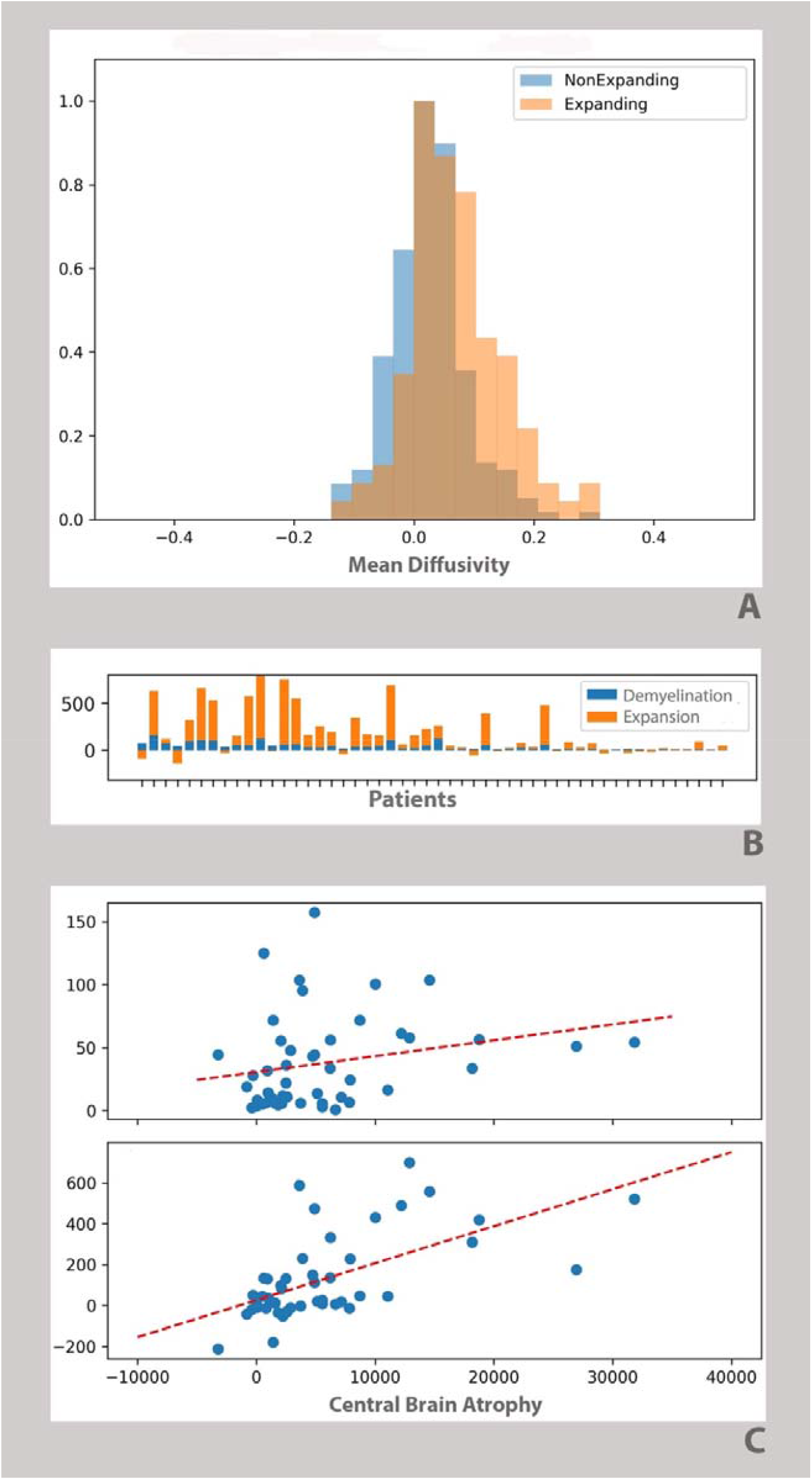
Effect of chronic demyelination and slow-burning inflammation on rarefication of chronic lesions: a. Normalised distribution of nMD change in individual voxels of the lesion core in stable and expanding lesion groups, Mean Diffusivity: μm^2^/ms b. Patient-wise modelling of the effect of permanent demyelination (blue) and low-grade inflammation at the lesion rim (orange) on rarefication of the tissue within lesional core. Vertical axis indicates equivalent of tissue loss-arbitrary units. c. Correlation of the tissue loss caused by chronic demyelination (top panel) and lesion expansion (bottom panel) with central brain atrophy. Vertical axis indicates equivalent of tissue loss-arbitrary units.

### Axonal loss caused by chronic demyelination of the optic nerve

To verify the level of axonal damage caused by the chronic demyelination, a subgroup of 15 patients with extensive unilateral chronic demyelination of the optic nerve following earlier optic neuritis (ON) was selected from the main study cohort. The group was followed for a median of 7.2 years (range 6.5-8 years). ON eyes demonstrated a significantly faster annual rate of RNFL thinning compared to fellow eyes (0.32% vs. 0.13 %, p=0.02, t-test).

Consequently, an accelerated loss of RGC axons in eyes exhibiting chronic demyelination of the optic nerve versus fellow (non-optic neuritis) eyes constituted an annual increase of 0.19% (0.32% - 0.13 %).

### Relative severity of axonal loss caused by chronic demyelination vs slow-burning inflammation

Assuming independent contributions of chronic demyelination and slow burning inflammation to the axonal loss (and to the associated tissue rarefication inside the chronic lesions), it is possible to estimate the relative weight of each factor on axonal attrition.

The core of chronic lesions is typically fully demyelinated[25], regardless of the degree of lesion expansion. Therefore, based on the analysis of stable lesions, chronic demyelination is likely to result in an annual loss of lesional axons equivalent to a 0.8% increase in nMD across 4 years of follow-up. Providing this reasoning is correct, the relative contribution of each factor to lesional tissue damage can be calculated in individual patients. Thus, assuming that the effect of chronic demyelination is equal in all demyelinated lesions, the damage caused by permanent loss of myelin during 4 year follow-up period will be proportional to the baseline lesion volume (i.e. MDdem=Vb x 0.8%, a where Vb represents baseline lesion volume and MDdem is proportion of MD change caused bu chronic demyelination). Furthermore, knowing the total change of tissue rarefication in each lesion during the same period (Vb x nMD change) we can calculate the effect of lesion expansion on tissue rarefication within the lesion (MDexp) by subtracting the change of MD related to chronic demyelination: MDexp=(Vb x nMDchange) - MDdem

Figure 6b, demonstrates the proportional contribution of chronic demyelination (blue) and low-grade inflammation at the lesion rim (orange) to progressive rarefication of lesional tissue, suggests the dominating role of the latter in the majority of patients. To verify the relative role of two mechanisms in axonal damage and subsequent degeneration, an equivalent of the tissue loss caused by each mechanism was correlated with central brain atrophy. The result of the correlation analysis also demonstrated that tissue damage caused by lesion expansion strongly linked to the loss of periventricular white matter (r=0.25, p=0.1 and r=0.61, p<0.001 for chronic demyelination and lesion expansion respectively). A similar result was demonstrated by the Linear Regression analysis of CBA (r-0.52), which showed a highly significant contribution of the lesion expansion component (Standardized Coef Beta=0.5, p=0.005), but not chronic demyelination (Standardized Coef Beta=0.1, p=0.5).

## Discussion

In the current study, we aimed to model the relative contribution of permanent demyelination and slow-burning inflammation at the rim of chronic lesions to progressive axonal loss in RRMS patients.

We demonstrated that while both mechanisms are likely to cause loss of white matter tissue inside the chronic lesions, slow-burning inflammation, when present, markedly exceeds the effect of chronic demyelination.

In this study, an increase in isotropic water diffusion (MD) within the core of chronic lesions was employed as an indicator of axonal loss. The use of MD as a marker for structural changes in white matter is supported by the strong correlation between MD levels and lesional T1 hypointensity, [3] an imaging metric closely related to the extent of tissue damage within MS lesions [16] [26]. This is also in line with previous combined MRI-histological[15] and DTI studies, which revealed the highest diffusivity in lesions with significant tissue loss [27] [28] [29] [30]

The elevation of MD, previously demonstrated in chronic white matter MS lesions, has been shown to reflect an enlargement of the extracellular space[31] [27] [32] [29] [30], caused by a combination of ongoing tissue loss and relatively rigid lesional structure (which is due to fibrillary gliosis developed during the acute stage of lesion formation [17] [33]), and its gradual increase has been associated with ongoing axonal loss.[3] [14]

Average degree of tissue loss calculated based of tissue rarefication (37%) is comparable with histopathological fundings in relapsing-remitting MS patients. [34] [24]

### MD dynamic range in white matter

To our knowledge, the relationship between MD and white matter damage inside MS lesions has not previously been quantified. While the best way to verify this relationship is to perform post-mortem studies correlating diffusion findings with histopathology of patients with MS, the results of such studies may not be directly applicable to *in vivo* human pathology since the water content changes dramatically depending on the post-mortem interval or brain fixation technique. [35] [36] [37]

Therefore, in order to model an association between isotropic water diffusion and loss of white matter tissue we first established the dynamic range of MD values in a substantial cohort of MS patients, which included various degrees of white matter abnormality ranging from unaltered (NAWM) to severely damaged (black holes) tissues.

The average value and narrow distribution of MD in NAWM, found in our study, are indicative of normal axonal density and consistent with previously published data [38].

Conversely, the distribution of MD in lesional voxels displayed a wide range of values from 0.75 μm^2^/ms to 2.0 μm^2^/ms with the largest detected MD elevation is likely signifying an extreme axonal damage typically seen in the core of destructive lesions [29] [27] [32] [29]. Considering that axonal loss in chronic white matter lesions can reach 90-100% [39] [40] [16] [24] the dynamic range of MD change between the average value in NAWM and the largest value in the core of chronic lesions is likely to cover the entire spectrum of axonal damage possible in white matter of RRMS patients (i.e. from 0% to 100%). Following this logic, and assuming that the degree of white matter rarefication within the dynamic range is proportional to the level of axonal attrition, it is plausible to infer that elevation of MD normalised within this interval represents a reasonably accurate measure of relative axonal loss in the lesion core during the follow-up period.

Using this approach, the rate of axonal attrition was calculated in expanding and stable chronic MS lesions. The rationale behind separating patients into two groups was as follows: while all chronic lesions (and the lesion core in particular) are completely free of myelin, [17] the same lesions also display slow-burning inflammation at their rims (as measured by their imaging equivalent, i.e. lesion expansion), providing, therefore, an additional source of axonal transection and subsequent degeneration. Hence, we hypothesised that whereas the alteration of MD in stable lesions is likely to indicate axonal attrition caused exclusively by the chronic demyelination, an additional increase of MD in expanding lesions is driven by the axonal transection caused by the slow-burning inflammation at the lesion rim.

### Axonal loss associated with chronic demyelination

Our results demonstrated a small, but significant increase in MD in the core of chronic stable lesions, which, based on the discussion above, is likely to reflect axonal attrition triggered by the permanent loss of myelin. Remarkably, a similar rate of accelerated axonal loss was also detected in severely demyelinated axons of the optic nerve using a different measuring modality. OCT, used in this analysis, allows measurement of RNFL thickness, which consists solely of RGC axons, providing, therefore, a direct and accurate assessment of axonal loss. More importantly, the analysis of RGC axonal loss in the visual system of patients with unilateral optic neuritis allows an inter-eye comparison of axonal loss between demyelinated and myelinated fibers (i.e. between optic neuritis and fellow eyes), which is not feasible in white matter of the brain. It is also worth noting that slow-burning inflammation is not typically observed in lesions of the optic nerve, making it a unique target for investigating the effect of chronic demyelination in isolation.[41]

The view that permanent demyelination may cause axonal damage by rendering axons vulnerable to physiological stress [42] [43] [44] by the way of increased energy demands on axonal conduction along demyelinated axons, leading to compromised axoplasmic ATP production[45], ionic imbalance and Ca^2+^-mediated axonal degeneration,[46] has recently been advocated based on experimental data. It was also suggested that lack of trophic support from myelin or oligodendrocytes and disruption of normal axon-myelin interactions together with the damaging effect of activated astrocytes may be implicated in accelerated degeneration of chronically demyelinated axons[43][44][47].

Our findings, which demonstrated an increase in tissue rarefication in stable MS lesions and RNFL thinning in eyes affected by extensive and long-standing demyelination, strongly support the notion that chronic loss of myelin does promote axonal loss. The magnitude of the axonal loss caused by permanent demyelination, however, appears to be very modest.

### Axonal loss associated with slow-burning inflammation

Conversely, in expanding lesions, the observed rate of MD elevation in the lesion core was markedly higher. The average rate of longitudinal tissue rarefication in this group was about 5-fold higher compared to the non-expanding group. Considering that the effect of chronic demyelination on axonal survival is likely to be similar regardless of lesion expansion (since all lesions are equally demyelinated), the magnitude of additional tissue rarefication in this group indicates that the impact of lesion expansion on lesional tissue damage is considerably more severe. The proportional contribution of the two mechanisms, however, varies considerably between patients from approximately equal to largely driven by lesion expansion, as the analysis of individual cases demonstrated. The effect of lesion expansion dominates (often dramatically) in at least half of the cases. Consequently, lesion expansion appears to be a major factor responsible for axonal damage and subsequent degeneration, as further supported by strong correlation with CBA and regression analysis. A significant positive correlation between the increase in diffusivity in the lesion core and the degree of lesion expansion, observed in this study, also strongly supports this notion.

Expanding lesions constituted about 25% of the total lesion population in our cohort, which is consistent with the current literature [48] As stated above, the expansion of MS lesions represents the imaging equivalent of slow-burning inflammation at the rim of some chronic lesions. [49][50]. Histopathologically, slowly expanding lesions constitute a recently classified subset of mixed active/inactive lesions (previously called smoldering lesions). [51]

[52] They are characterised by a narrow rim of macrophages/microglia containing MBP+ or PLP+ myelin degradation products, which reflects an ongoing low-grade demyelinating activity. Such lesions typically reveal an inactive hypocellular and completely demyelinated plaque core that is surrounded by a rim of profound microglial activation, ongoing smoldering demyelination and axonal injury.[53] [54] [55] [56] Compared to the cellular composition at the rim of chronic inactive lesions (which predominantly show oligodendrocytes), the edges of chronic active lesions feature significantly more pro-inflammatory immune cells.[57] This low-grade inflammation at the edge of chronic lesions has been implicated as one of the main forces driving MS progression, including neurodegeneration, brain atrophy and disability worsening.[49] [58] [6] [50] [59]

This is also in agreement with pathological studies demonstrating that active demyelination in mixed active/inactive lesions correlates with axonal injury in these lesions. [60] [61]

An association between the rate of chronic lesion expansion and increase of tissue rarefication inside the lesions (suggesting that ongoing low-grade inflammation promotes axonal injury or transection at the lesion edge and subsequent progressive tissue damage within the lesion) has been recently observed by several groups [6] [58] [62]. This relationship has been attributed to Wallerian and retrograde degeneration, which leads not only to loss of extra-lesional section of the transected axons (and, as a result, to brain atrophy), but also to attrition of the intra-lesional part of those axons which traverse the lesion itself (and, as a result, increase rarefication of lesional tissue). [10]

It is likely that some axons transected within newly appearing (acute) lesions also traverse chronic lesions analysed in this study. Therefore, to minimize the potential effect of Wallerian and retrograde degeneration of those axons on tissue rarefication inside the chronic lesions (and, consequently, on progressive change of MD), all voxels containing streamlines intersecting with new brain lesions were excluded from the longitudinal analysis of diffusivity.

This study has several limitations. No direct quantitative histological validation of MD as a measure of axonal attrition has been performed. Assumptions of proportional relationship between axonal loss and MD elevation as well as additive nature of the effect of chronic demyelination and slow-burning inflammation on axonal loss are not yet experimentally confirmed. Furthermore, Wallerian degeneration caused by acute spinal or cortical lesions can also potentially contribute to tissue rarefication in chronic white matter lesions. Because of the small size of treatment groups and lesion-based, rather than patient-based analysis, the effect of individual therapy on study outcomes was not assessed.

In conclusion, the current study demonstrated a significant increase in water content in chronic MS lesions. The degree of progressive tissue rarefication, however, is markedly higher in slowly expanding compared to stable lesions. This suggests that slow-burning inflammation at the lesion rim, when present, is likely to play a more significant role in axonal attrition than chronic demyelination.

## Data Availability

All data produced in the present study are available upon reasonable request to the authors

